# Survival benefits of cytoreductive nephrectomy in patients with metastatic renal cell carcinoma: evidence from a SEER-based retrospective cohort study

**DOI:** 10.1101/2025.01.28.25321237

**Authors:** Xiongwu Peng, Lingxing Duan, Runlin Shi

## Abstract

**Background:** Metastatic renal cell carcinoma (mRCC) is associated with poor prognosis, with a 5-year survival rate of less than 15%. Cytoreductive nephrectomy (CN) has historically played a critical role in mRCC management, potentially enhancing systemic therapy efficacy by reducing tumor burden. However, its relevance in the era of targeted therapies and immune checkpoint inhibitors (ICIs) has been questioned.

**Objective:** This study evaluates the survival benefits of CN in mRCC patients using real-world, population-based data from the SEER database.

**Methods:** A retrospective cohort analysis of 6,030 mRCC patients was performed using data from 2010 to 2017. Propensity score matching (PSM) minimized selection bias, yielding 1,350 matched patients. Kaplan-Meier survival curves and multivariate Cox proportional hazards models assessed the impact of CN on overall survival (OS) and RCC-specific survival (CSS), stratified by demographic and clinical characteristics.

**Results:** CN was associated with a 71% reduction in all-cause mortality (HR = 0.29, 95% CI = 0.25–0.33) and RCC-specific mortality (HR = 0.29, 95% CI = 0.25–0.34). Five-year OS rates were 31.5% in the CN group versus 3.6% in the non-CN group. Survival benefits were consistent across subgroups, including patients with high-grade or advanced-stage tumors, underscoring the role of CN within multimodal treatment strategies.

**Conclusion:** CN confers significant survival advantages in mRCC, even in challenging clinical scenarios. These findings reinforce the importance of integrating CN into multimodal therapeutic frameworks, particularly alongside modern systemic therapies. Further prospective studies are warranted to optimize patient selection and treatment sequencing.

## Introduction

Renal cell carcinoma (RCC) accounts for 3%–5% of global malignancies(1). It represents the most prevalent form of kidney cancer, representing nearly 90% of all renal neoplasms(2). Despite advancements in early detection and therapeutic interventions, a substantial proportion of patients are diagnosed with metastatic disease (M1 stage), which is linked to a five-year survival rate of under 15%(3, 4). These statistics underscore the pressing need to develop more effective treatment strategies to improve outcomes for this vulnerable patient population.

Cytoreductive nephrectomy (CN), a surgical intervention targeting the removal of the primary tumor, has traditionally been a key component in the treatment of metastatic renal cell carcinoma (mRCC)(5). The procedure is hypothesized to alleviate tumor burden, potentiate the host immune response, and augment the efficacy of subsequent systemic therapies(6). However, with the rise of targeted therapies like tyrosine kinase inhibitors (TKIs) and immune checkpoint inhibitors (ICIs), the significance of CN has come under growing scrutiny(7–9). Landmark randomized trials, including CARMENA and SURTIME, have cast doubt on the universal benefit of CN, particularly in patients with poor performance status or extensive metastatic burden(8–12). Nevertheless, the applicability of these findings to real-world populations remains constrained by the trials’ restrictive inclusion criteria and limited sample sizes(13). Consequently, the optimal patient subgroups most likely to derive survival benefit from CN in the modern era of systemic therapies remain inadequately defined(8).

Given these uncertainties, we postulate that CN retains the potential to confer substantial survival benefits in carefully selected patients with mRCC, especially when integrated into a multimodal therapeutic framework. To test this hypothesis, we analyzed data from the Surveillance, Epidemiology, and End Results (SEER) database, a comprehensive population-based cancer registry. Our study design entailed meticulous patient selection, extensive stratification, and rigorous statistical analyses to evaluate the impact of CN on overall survival (OS) and RCC-specific survival (RCC-SS).

In addition, this investigation employed stratified analyses across sociodemographic, clinical, and pathological characteristics to rigorously evaluate the potential survival advantages conferred by CN. Kaplan-Meier survival curves and Cox proportional hazards models revealed significantly lower overall and RCC-specific mortality among patients undergoing CN compared to those receiving non-surgical management. Furthermore, stratified analyses demonstrated a remarkable consistency in survival benefits across diverse clinical subgroups, underscoring the integral role of CN within multimodal treatment paradigms.

By employing a propensity score-matched cohort and leveraging advanced statistical methodologies, including Cox proportional hazards and Fine and Gray competing risks models, we identified critical patient subgroups most likely to benefit from CN. This research provides important insights into the current role of CN in clinical practice, helping to refine therapeutic approaches for patients with mRCC. Ultimately, our findings emphasize the imperative to identify patient-specific factors to maximize the therapeutic efficacy of CN and improve long-term outcomes in this challenging disease landscape.

## Methods

### Data source

The SEER database, utilized in this analysis, includes data from 17 U.S. states between 2000 and 2019, as per its 2021 edition. Covering approximately 26.5% of the U.S. population, it provides a comprehensive resource for examining cancer incidence, survival, and treatment patterns. Its geographic and demographic diversity, coupled with rigorous data standards, ensures robust insights into cancer trends and disparities.

The Ethics Committee of the Third People’s Hospital of Pingxiang City granted an exemption for this study, as the SEER program solely utilizes anonymized, population-based cancer registry data. Since the database is publicly available and does not require direct engagement with human subjects, formal ethical approval was considered redundant.

### Cohort selection

The case list was generated using SEER*Stat software, version 8.4.4, and included patients diagnosed with metastatic (M1 stage) renal parenchymal carcinoma, classified under ICD-O code C64.9. The dataset captured an extensive array of variables, encompassing demographic factors (age, sex, ethnicity, marital status), clinical characteristics (year of diagnosis, tumor grade, laterality, histologic subtype, T and N stages), and treatment details (radiation therapy and chemotherapy). This comprehensive dataset forms a robust foundation for analyzing disease patterns, treatment strategies, and outcomes in metastatic renal cell carcinoma.

To refine the cohort, 124,007 cases diagnosed between 2000–2009 and 2018–2019 were excluded, restricting the analysis to diagnoses made between 2010 and 2017. Patients under the age of 18 were further excluded, resulting in an initial cohort of 91,842 patients. Subsequently, cases with missing T and N staging, patients who had received chemotherapy or radiotherapy, and those who had not undergone partial nephrectomy (PN) or radical nephrectomy (RN) were excluded, eliminating 7,610 cases. Additionally, cases with incomplete outcome data and those diagnosed with renal pelvis cancer (International Classification of Diseases for Oncology code C65.9) were removed, further reducing the cohort by 96 cases. This rigorous selection process yielded a final cohort of 6,030 eligible patients.

To address potential selection bias, propensity score matching (PSM) was employed, resulting in a matched cohort of 1,350 patients for subsequent analysis.

## Vital Status

Patient status at the latest follow-up was assessed using the SEER9 “cause of death (COD) to site recode” variable, allowing precise classification of survival outcomes. Patients were categorized as alive at the most recent follow-up, deceased due to renal cell carcinoma, or deceased from non-renal causes. The primary endpoint of the analysis was overall mortality, providing a comprehensive measure of survival. Secondary outcomes included renal cancer-specific mortality and deaths attributable to non-renal causes, offering critical insights into the interplay of disease progression and competing health risks.

Temporal data were derived from the “months of survival” variable, which spanned from the date of diagnosis to the most recent follow-up. Survival duration, expressed in months, was computed using SEER*Stat software by subtracting the diagnosis date from the last contact date (or study endpoint). The calculation employed a conversion factor of 365.24 days per year divided by 12 to approximate the number of days in a month. The study endpoint was set as December 31, 2019.

### Balancing Cohorts Using Propensity Score Matching

Propensity scores were calculated using logistic regression, incorporating variables such as demographics (age, sex, race), tumor characteristics (T stage, N stage, laterality, histologic subtype, grade), sequence number, and tumor count to adjust for treatment selection bias. A one-to-one matched cohort was then created using the nearest-neighbor method with a caliper width of 0.001, ensuring precise pairing and balance between surgical and non-surgical groups for outcome comparison.

### Statistical Analysis

Descriptive analyses were conducted for all variables, with patients categorized according to the treatment modality they received, classified as either surgical or non-surgical. Non-surgical management was defined as the absence of surgery, while surgical intervention included only partial nephrectomy (PN) or radical nephrectomy (RN). OS was defined as the period from the date of RCC diagnosis to death from any cause, while RCC-SS was the duration from diagnosis to death specifically due to RCC.

Univariate (unadjusted) analyses were performed to identify covariates associated with mortality, and stratified (adjusted) analyses were employed to evaluate the differential impact of treatment within specific population subgroups. KM curves were used to construct survival curves for patients with RCC, stratified by treatment type. KM curves for all-cause mortality and RCC-specific survival, further stratified by T and N stages, were generated to examine treatment effects across various subgroups.

The Cox proportional hazards model was employed to evaluate the influence of various factors, including age, tumor laterality, race, tumor grade, year of diagnosis, histologic subtype, sex, T and N stages, treatment modality, sequence number, tumor burden, and marital status, on both overall mortality and RCC-SS. Furthermore, the cumulative incidence of competing risks was utilized to estimate RCC-specific mortality, while appropriately accounting for the impact of competing events. All statistical analyses were conducted using Empower (R) (www.empowerstats.com, X&Y Solutions, Inc., Boston, MA, USA) and R version 3.6.3 (http://www.R-project.org), both of which are recognized for their robust data processing capabilities and extensive analytical features. Statistical significance was considered achieved when the p-value was less than 0.05.

## Results

### Baseline Characteristics of the Patient Population

Of the entire cohort, only 1,560 patients (25.9%) underwent surgical resection of the primary tumor, whereas the majority (4,471 patients, 74.1%) did not receive any surgical intervention. Patients who underwent surgery were disproportionately older, white, married, and diagnosed with clear cell carcinoma, suggesting that both demographic characteristics and tumor-specific features exerted considerable influence over treatment decisions. Moreover, the choice between surgical and non-surgical management was strongly associated with tumor stage and grade. In particular, among patients with T3 stage tumors, only 16.09% did not undergo surgical resection, underscoring the pivotal role of surgical intervention in the management of advanced-stage disease.

To minimize potential confounding factors and enable robust comparisons, a propensity score-matched cohort comprising 1,350 patients was established. Of these, 655 individuals were allocated to the surgical group, while 695 were assigned to the non-surgical group. Following the matching process, the baseline clinical characteristics between the two cohorts were largely comparable, with no significant differences observed, except for a few variables such as sex, tumor grade, laterality, and T stage. This methodologically rigorous approach substantially mitigated bias, thereby enabling a more reliable and nuanced evaluation of treatment outcomes between the surgical and non-surgical cohorts.

### Univariate analysis

The unadjusted analysis revealed a significant survival advantage for the surgical cohort, with lower all-cause mortality (hazard ratio [HR] = 0.40) and renal cell carcinoma-specific mortality (HR = 0.30) compared to those in the non-surgical group. These results were highly statistically significant (p < 0.0001), suggesting that surgical resection is associated with improved overall survival and RCC-specific survival.

Further investigation into the factors influencing survival outcomes highlighted the complexity of the disease, as both all-cause and RCC-SS were influenced by multiple clinical and demographic variables, including marital status, tumor grade, histologic subtype, T stage, and N stage. The multivariate regression analysis, adjusted for these factors, reinforced the survival benefits of surgery across different patient subgroups.

### Stratified analysis

The stratified adjusted analysis (Supplementary Table S1) provides a detailed examination of the influence of CN on survival outcomes in mRCC patients, taking into account variables such as age, sex, race, tumor grade, histologic subtype, and staging. The results indicate that patients who underwent CN exhibited a significant reduction in both all-cause mortality and kidney cancer-specific mortality across various subgroups.

Notably, younger patients (40–49 years: HR = 0.34, p < 0.0001), both males (HR = 0.38, p < 0.0001) and females (HR = 0.35, p < 0.0001), White individuals (HR = 0.35, p < 0.0001), and those with clear cell histology (HR = 0.33, p < 0.0001) derived the most pronounced survival benefits. Patients with lower-stage tumors (T1–T3) and lower-grade lesions (G1–G2) exhibited the greatest advantage (e.g., G2: HR = 0.17, p < 0.0001). Although patients with advanced-stage (T4) or high-grade tumors demonstrated comparatively smaller benefits, the survival advantage conferred by CN remained statistically significant.

Collectively, these results underscore the pivotal role of CN within multimodal therapeutic strategies, highlighting its substantial survival benefits in specific, well-defined patient subgroups.

### Multivariate analysis

In our multivariate regression analysis, patients who underwent surgical resection demonstrated substantially lower rates of both OS and RCC-SS compared to their non-surgical counterparts (Table 3). When evaluating OS, the surgical cohort exhibited a markedly reduced risk of mortality across all models, including the unadjusted analysis and Models I and II, which incorporated adjustments for sociodemographic and clinical variables. Notably, the hazard ratio (HR) for OS among patients who underwent surgery was 0.37 (95% CI = 0.37–0.42, p < 0.0001), corresponding to a 63% reduction in mortality risk. This survival benefit remained statistically robust throughout all analytical frameworks, underscoring the pivotal role of surgical intervention in enhancing long-term outcomes.

**Table 1.**
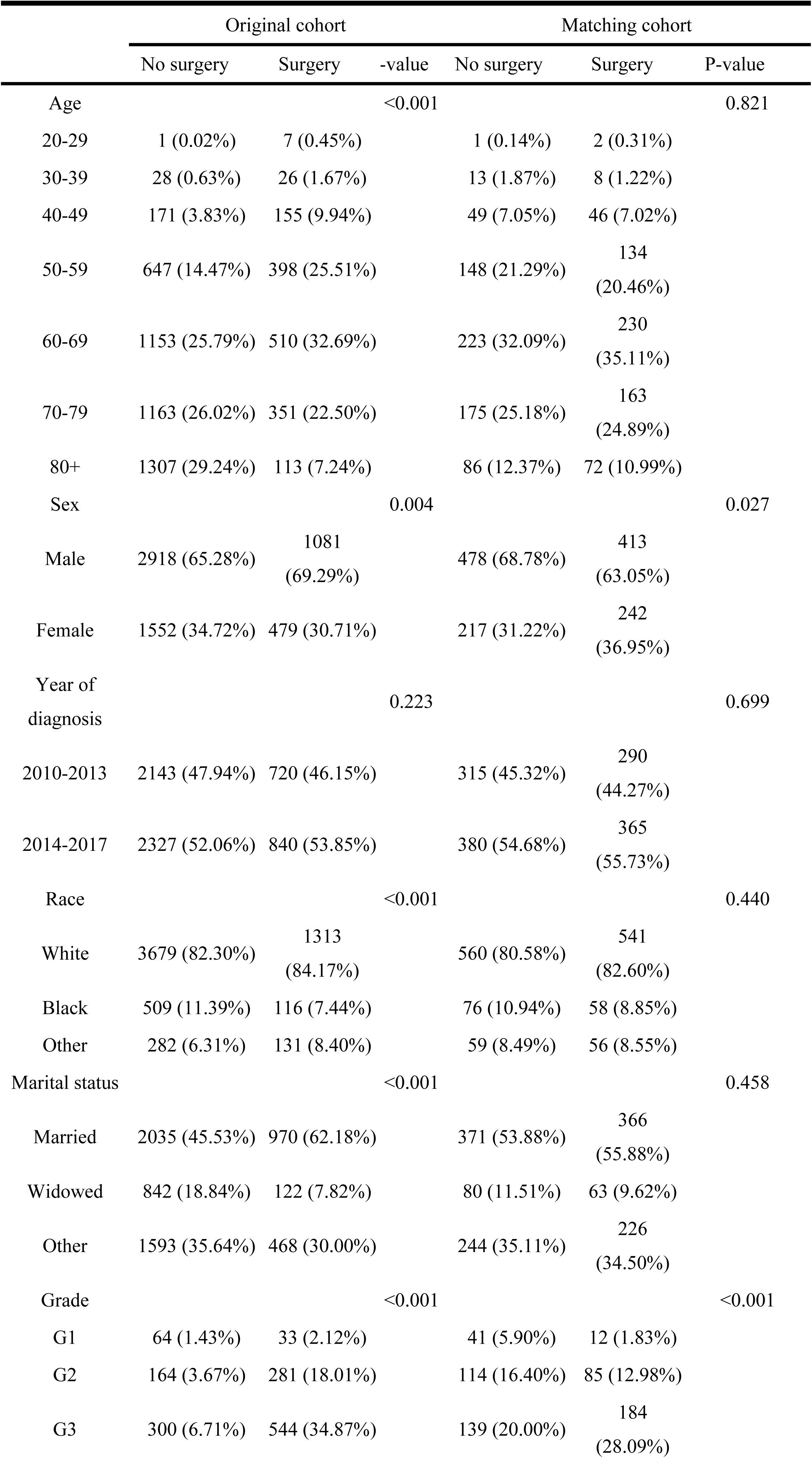
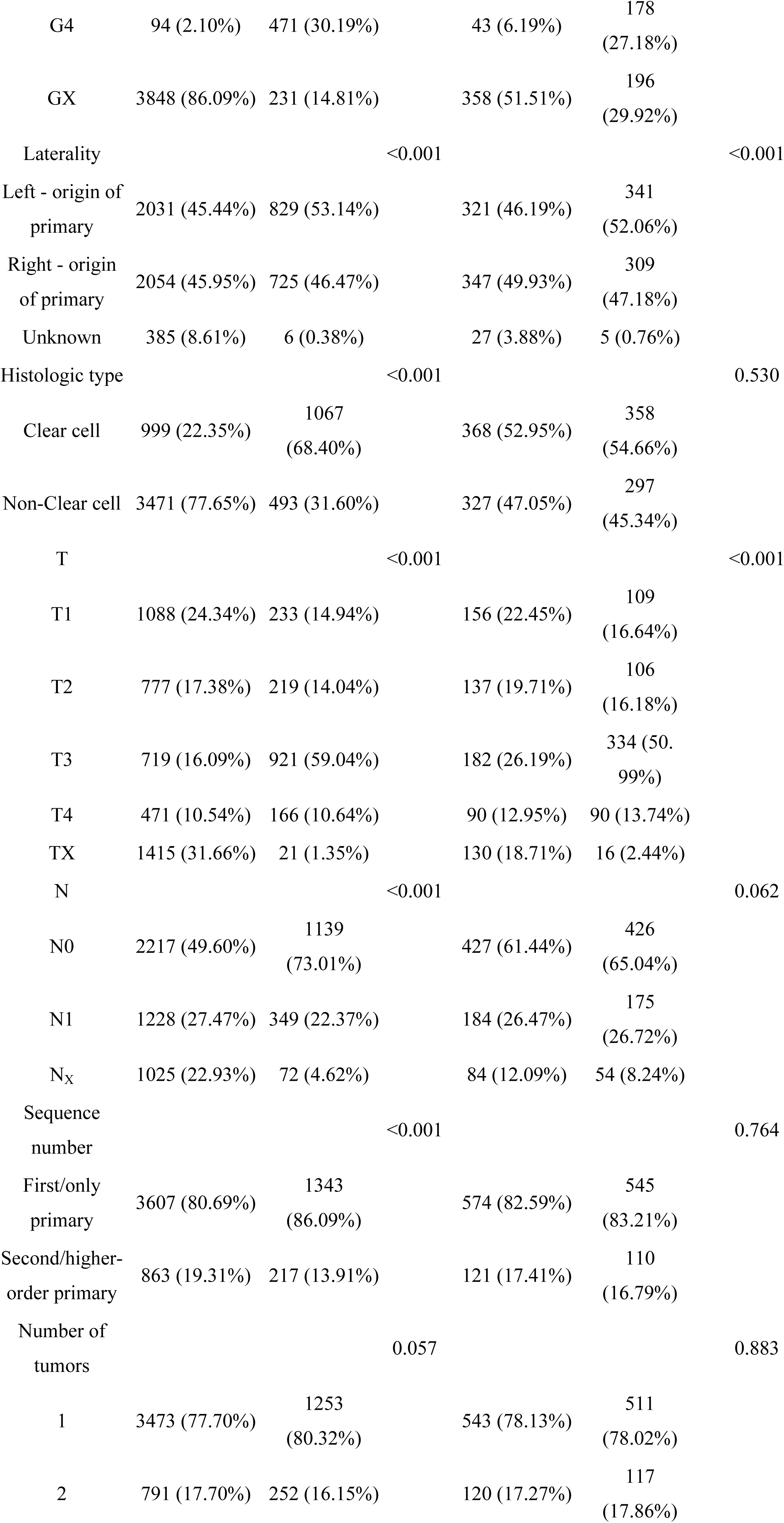
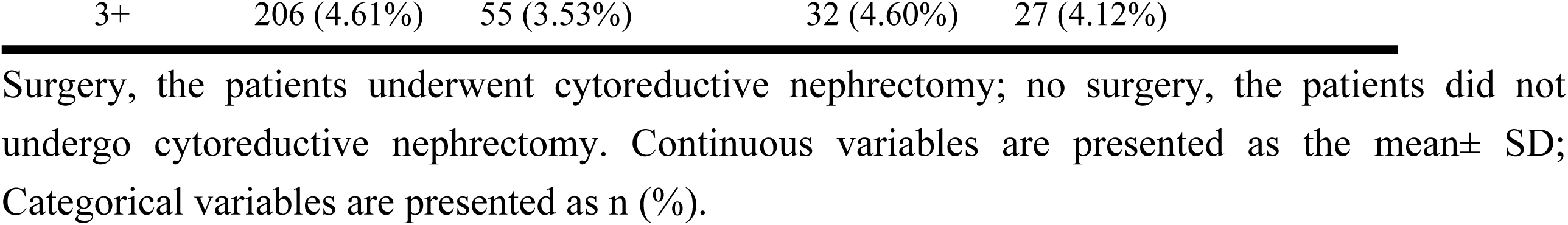
Baseline Characteristics of the Patient Population.

**Table 2.** Univariate analysis.

| Exposure | Statistics | Hazard Ratio(95%CI) p-value |  |
| --- | --- | --- | --- |
|  |  | Kidney cancer-specific mortality | All-cause mortality |
| Age |  |  |  |
| 20-29 | 3 (0.22%) | 1.0 | 1.0 |
| 30-39 | 21 (1.56%) | 0.73 (0.21, 2.51) 0.6187 | 0.76 (0.22, 2.60) 0.6657 |
| 40-49 | 95 (7.04%) | 0.61 (0.19, 1.95) 0.4081 | 0.65 (0.20, 2.06) 0.4630 |
| 50-59 | 282 (20.89%) | 0.63 (0.20, 1.98) 0.4304 | 0.67 (0.22, 2.10) 0.4953 |
| 60-69 | 453 (33.56%) | 0.66 (0.21, 2.07) 0.4778 | 0.72 (0.23, 2.24) 0.5691 |
| 70-79 | 338 (25.04%) | 0.67 (0.21, 2.10) 0.4939 | 0.77 (0.25, 2.40) 0.6485 |
| 80+ | 158 (11.70%) | 0.82 (0.26, 2.59) 0.7407 | 1.04 (0.33, 3.28) 0.9418 |
| Sex |  |  |  |
| Male | 891 (66.00%) | 1.0 | 1.0 |
| Female | 459 (34.00%) | 0.97 (0.85, 1.11) 0.6664 | 0.97 (0.86, 1.10) 0.6750 |
| Year of diagnosis |  |  |  |
| 2010-2013 | 605 (44.81%) | 1.0 | 1.0 |
| 2014-2017 | 745 (55.19%) | 0.87 (0.77, 0.99) 0.0391 | 0.90 (0.80, 1.02) 0.0961 |
| Race |  |  |  |
| White | 1101 (81.56%) | 1.0 | 1.0 |
| Black | 134 (9.93%) | 1.18 (0.96, 1.45) 0.1156 | 1.19 (0.98, 1.45) 0.0732 |
| Other | 115 (8.52%) | 0.98 (0.78, 1.24) 0.8846 | 0.97 (0.78, 1.20) 0.7765 |
| Marital status |  |  |  |
| Married | 737 (54.59%) | 1.0 | 1.0 |
| Widowed | 143 (10.59%) | 1.40 (1.14, 1.72) 0.0013 | 1.41 (1.16, 1.71) 0.0005 |
| Other | 470 (34.81%) | 1.03 (0.90, 1.18) 0.6913 | 1.02 (0.90, 1.16) 0.7249 |
| Grade |  |  |  |
| G1 | 53 (3.93%) | 1.0 | 1.0 |
| G2 | 199 (14.74%) | 0.69 (0.48, 1.00) 0.0482 | 0.69 (0.49, 0.96) 0.0289 |
| G3 | 323 (23.93%) | 0.97 (0.69, 1.37) 0.8600 | 0.94 (0.69, 1.29) 0.6948 |
| G4 | 221 (16.37%) | 1.18 (0.83, 1.68) 0.3443 | 1.02 (0.74, 1.41) 0.9088 |
| GX | 554 (41.04%) | 1.08 (0.78, 1.51) 0.6388 | 1.02 (0.75, 1.38) 0.9154 |
| Laterality |  |  |  |
| Left - origin of primary | 662 (49.04%) | 1.0 | 1.0 |
| Right - origin of primary | 656 (48.59%) | 1.03 (0.90, 1.17) 0.6823 | 1.00 (0.88, 1.12) 0.9433 |
| 3 | 32 (2.37%) | 1.08 (0.71, 1.64) 0.7262 | 1.21 (0.84, 1.75) 0.3015 |
| Histologic type |  |  |  |
| Clear cell | 726 (53.78%) | 1.0 | 1.0 |
| Non-Clear cell | 624 (46.22%) | 1.47 (1.29, 1.67) <0.0001 | 1.49 (1.32, 1.67) <0.0001 |
| T |  |  |  |
| T1 | 265 (19.63%) | 1.0 | 1.0 |
| T2 | 243 (18.00%) | 1.34 (1.07, 1.68) 0.0102 | 1.12 (0.91, 1.37) 0.2854 |
| T3 | 516 (38.22%) | 1.40 (1.16, 1.70) 0.0006 | 1.17 (0.98, 1.39) 0.0779 |
| T4 | 180 (13.33%) | 2.42 (1.93, 3.05) <0.0001 | 1.97 (1.59, 2.42) <0.0001 |
| TX | 146 (10.81%) | 1.92 (1.50, 2.46) <0.0001 | 1.68 (1.34, 2.10) <0.0001 |
| N |  |  |  |
| N0 | 853 (63.19%) | 1.0 | 1.0 |
| N1 | 359 (26.59%) | 1.79 (1.55, 2.07) <0.0001 | 1.73 (1.51, 1.98) <0.0001 |
| NX | 138 (10.22%) | 1.27 (1.03, 1.57) 0.0282 | 1.26 (1.03, 1.54) 0.0218 |
| Treatment |  |  |  |
| No surgery | 695 (51.48%) | 1.0 | 1.0 |
| Surgery | 655 (48.52%) | 0.37 (0.32, 0.42) <0.0001 | 0.36 (0.32, 0.41) <0.0001 |
| Sequence number |  |  |  |
| First/only primary | 1119<br>(82.89%) | 1.0 | 1.0 |
| Second/higher-order<br>primary | 231 (17.11%) | 0.80 (0.67, 0.95) 0.0120 | 0.94 (0.80, 1.10) 0.4141 |
| Number of tumors |  |  |  |
| 1 | 1054<br>(78.07%) | 1.0 | 1.0 |
| 2 | 237 (17.56%) | 0.78 (0.66, 0.93) 0.0065 | 0.92 (0.78, 1.07) 0.2765 |
| 3+ | 59 (4.37%) | 0.61 (0.42, 0.88) 0.0085 | 0.90 (0.67, 1.21) 0.4868 |

**Table 3.**
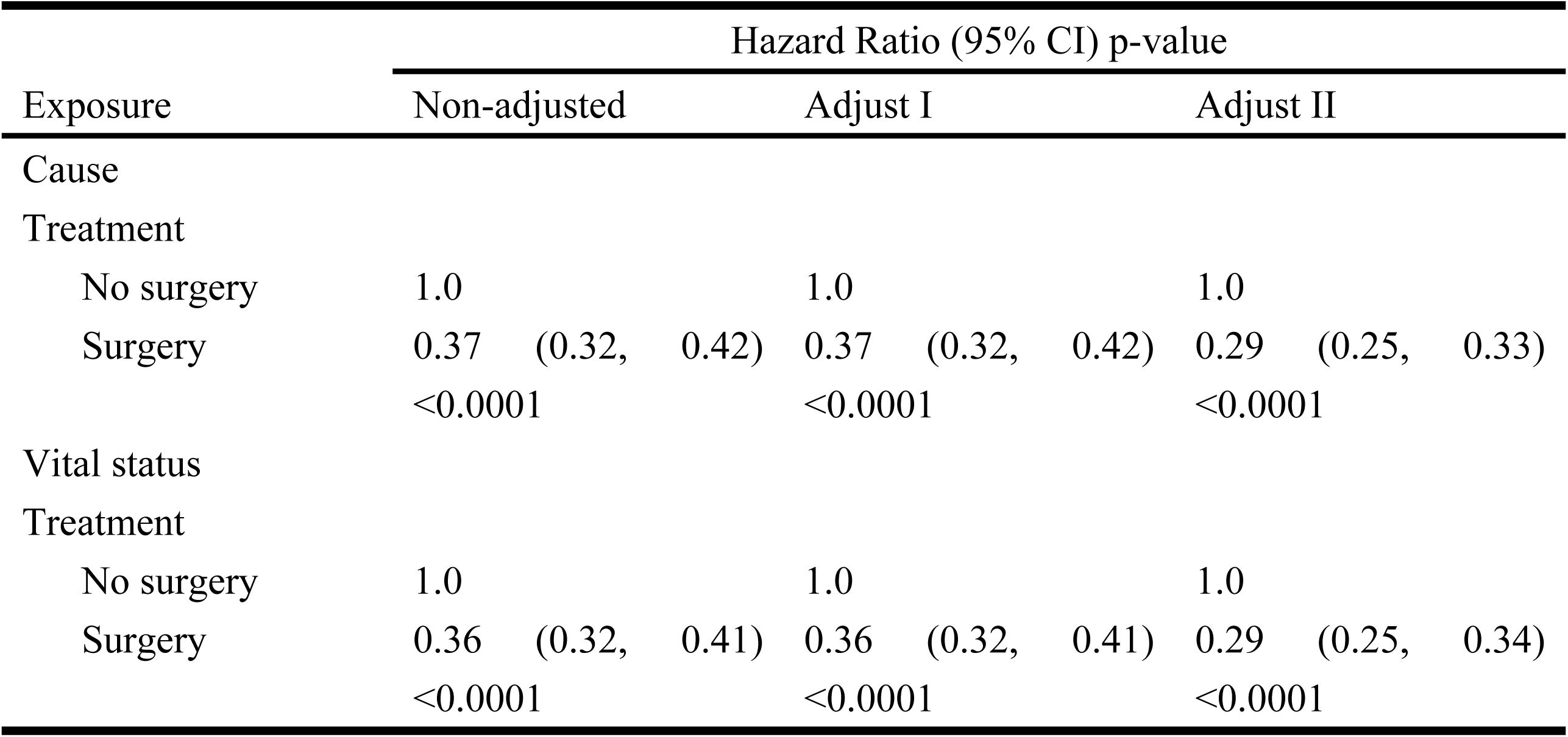
Multivariate analysis.

Similarly, the analysis of RCC-specific survival highlighted a consistent and significant benefit for patients who underwent surgical resection. Across both unadjusted and adjusted models, the risk of RCC-SS was reduced by 64% in the surgical cohort (HR = 0.36, 95% CI = 0.32–0.41, p < 0.0001). This benefit was even more pronounced in the fully adjusted model, which accounted for an extensive array of sociodemographic and clinical confounders, yielding an HR of 0.29 (95% CI = 0.25–0.34, p < 0.0001). This represents a remarkable 71% reduction in RCC-specific mortality, further emphasizing the significant protective effect of surgical treatment.

### OS and RCC-SS across treatment modalities

Patients who underwent surgical intervention demonstrated markedly superior survival outcomes compared to those who did not undergo surgery (Table 4). For overall survival (OS), the 1-year, 3-year, and 5-year survival rates were 60.97%, 41.79%, and 31.50%, respectively, in the surgery group, as opposed to 23.96%, 8.05%, and 3.56% in the non-surgery group. Similarly, RCC-SS at 1, 3, and 5 years was 63.87%, 46.23%, and 36.20% for the surgery group, compared to 28.40%, 11.47%, and 5.77% for the non-surgery group. These findings underscore the significant survival advantage conferred by surgical intervention.

**Table 4.** Overall survival and renal cancer-specific survival.

| Treatment | No surgery | Surgery |
| --- | --- | --- |
| Overall Survival |  |  |
| N | 695 | 695 |
| 1-year survival (95% CI) | 23.96% (21.17%–27.12%) | 60.97% (57.76%–64.37%) |
| 3-year survival (95% CI) | 8.05% (6.41%–10.11%) | 41.79% (38.27%–45.64%) |
| 5-year survival (95% CI) | 3.56% (2.53%–5.01%) | 31.50% (27.95%–35.50%) |
| Kidney cancer-special survival |  |  |
| N | 3795 | 3733 |
| 1-year survival (95% CI) | 28.40% (25.29%–31.90%) | 63.87% (60.63%–67.27%) |
| 3-year survival (95% CI) | 11.47% (9.31%–14.13%) | 46.23% (42.57%–50.21%) |
| 5-year survival (95% CI) | 5.77% (4.22%–7.88%) | 36.20% (32.36%–40.49%) |

CI, Confidence interval.

As illustrated in Table 4 and Figure 2A, patients in the surgery group achieved significantly higher OS compared to their non-surgical counterparts (p < 0.0001). The KM survival curve analysis corroborates these results, providing robust evidence of the survival benefit observed in the tabulated data.

**Figure 1.**
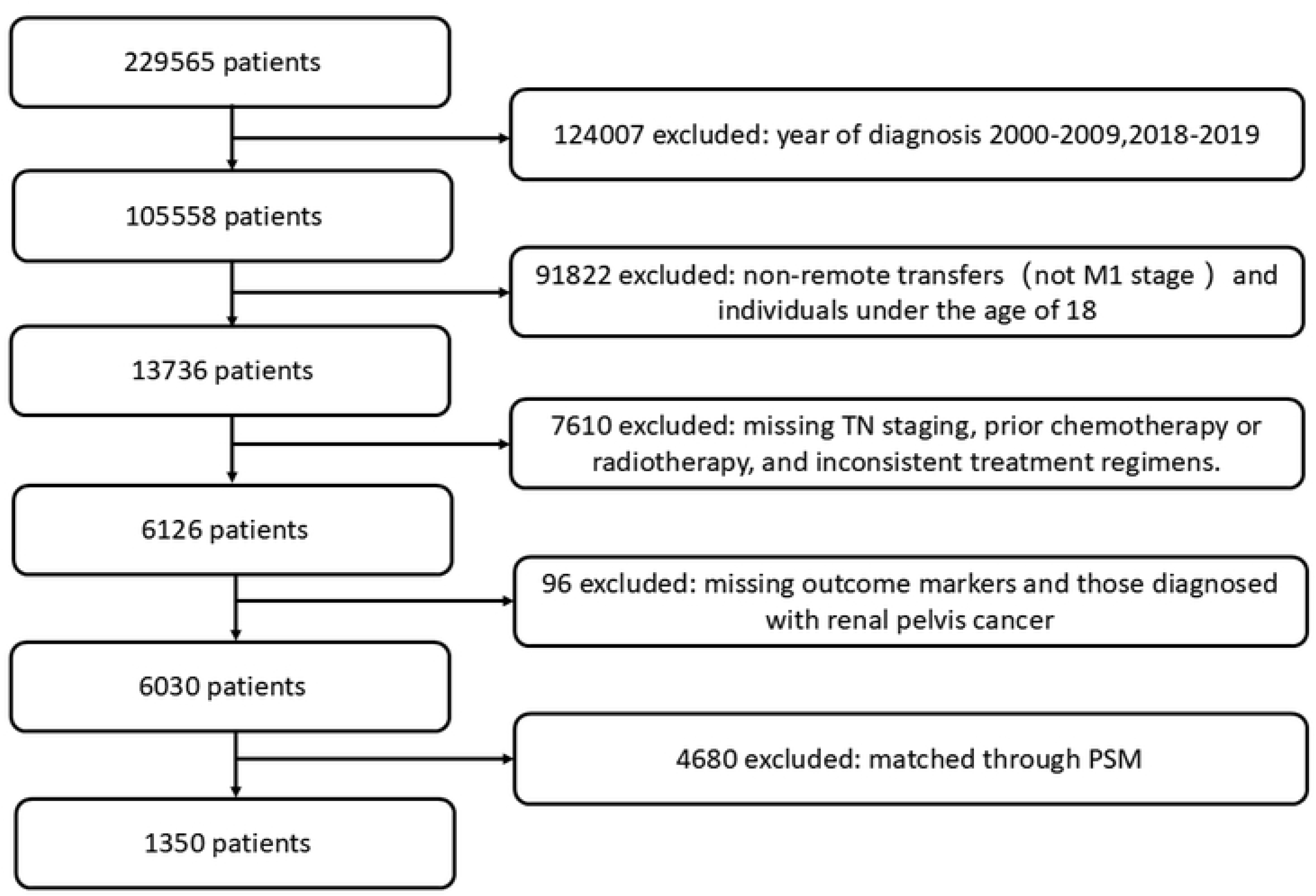
Flow chart used to select participants.

**Figure 2.**
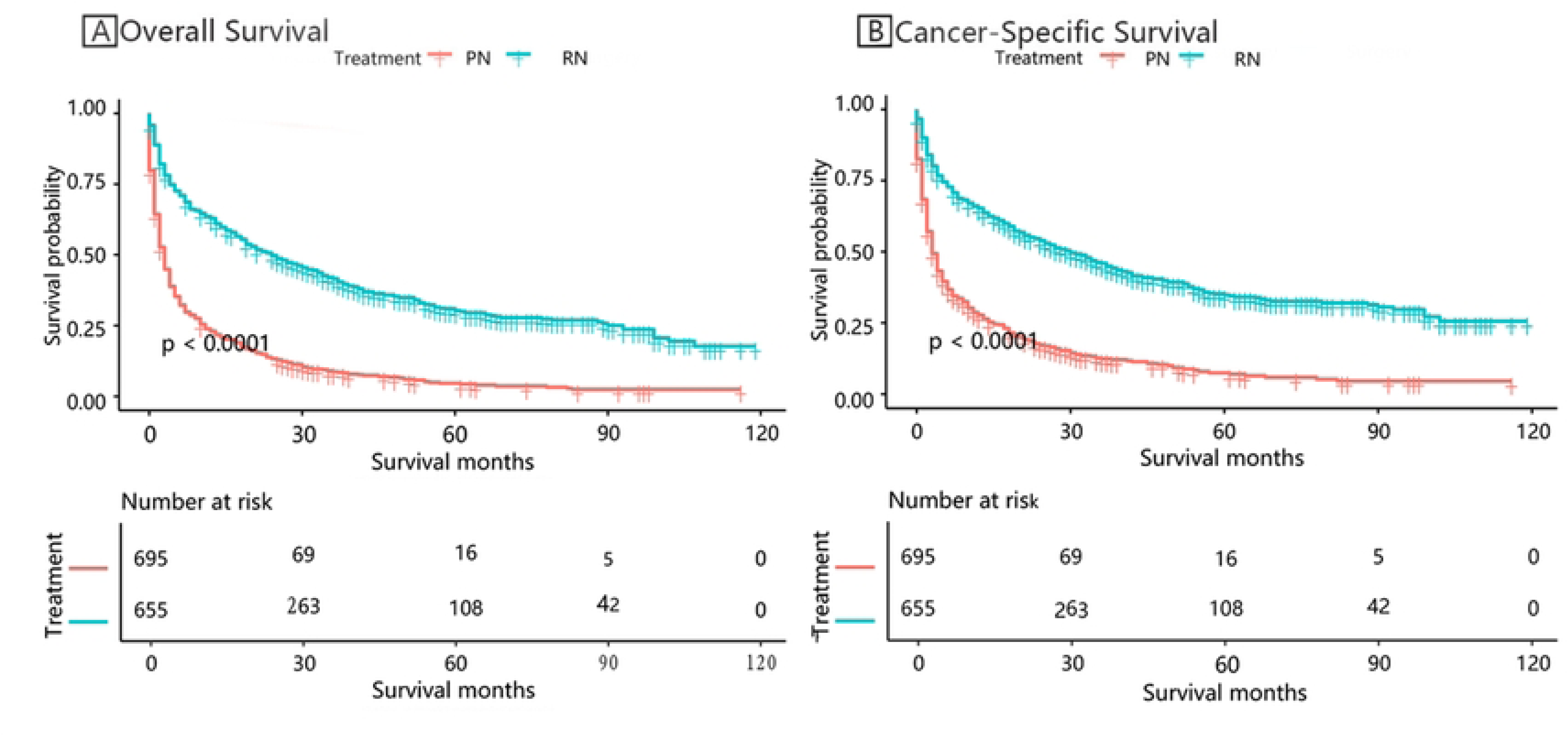
Survival outcomes stratified by treatment modalities.

In a similar fashion, RCC-SS, as presented in Table 4 and Figure 2B, was notably higher among patients who underwent surgery compared to those who did not (p < 0.0001). The 1-year, 3-year, and 5-year RCC-SS rates in the surgery group were 63.87%, 46.23%, and 36.20%, respectively, whereas in the non-surgery group, the corresponding rates were 28.40%, 11.47%, and 5.77%. This pattern was further validated by KM survival curve curve analysis, which consistently revealed a marked survival benefit favoring those who received surgical treatment.

Figures 3 and 4 further highlight this survival benefit, with OS and RCC-SS consistently favoring the surgery group across all stratifications. Statistically significant differences were observed (p < 0.05), reaffirming the critical role of surgical intervention in improving both OS and RCC-SS outcomes.

**Figure 3.**
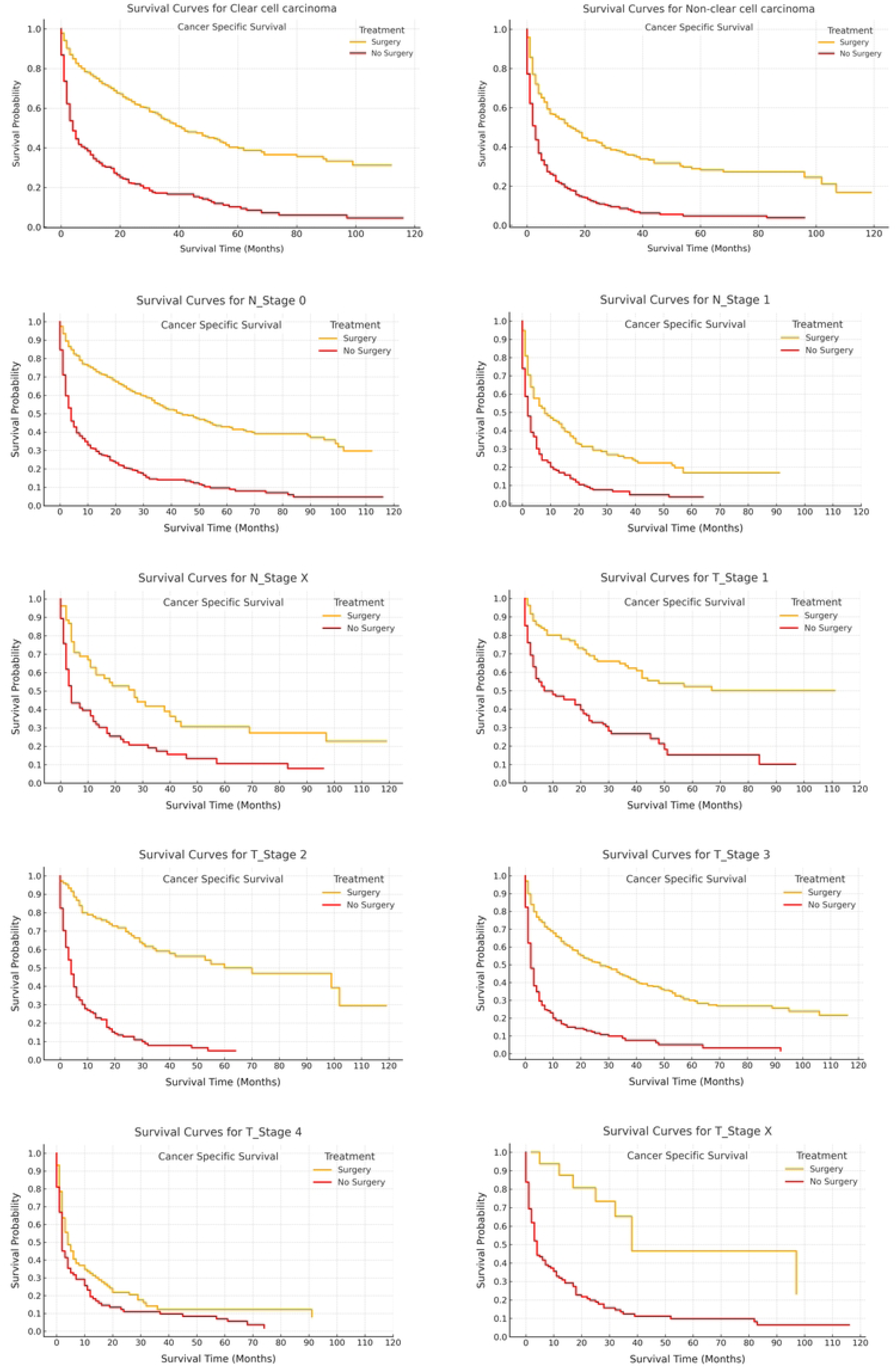
Kidney cancer-specific survival stratified by treatment modalities.

**Figure 4.**
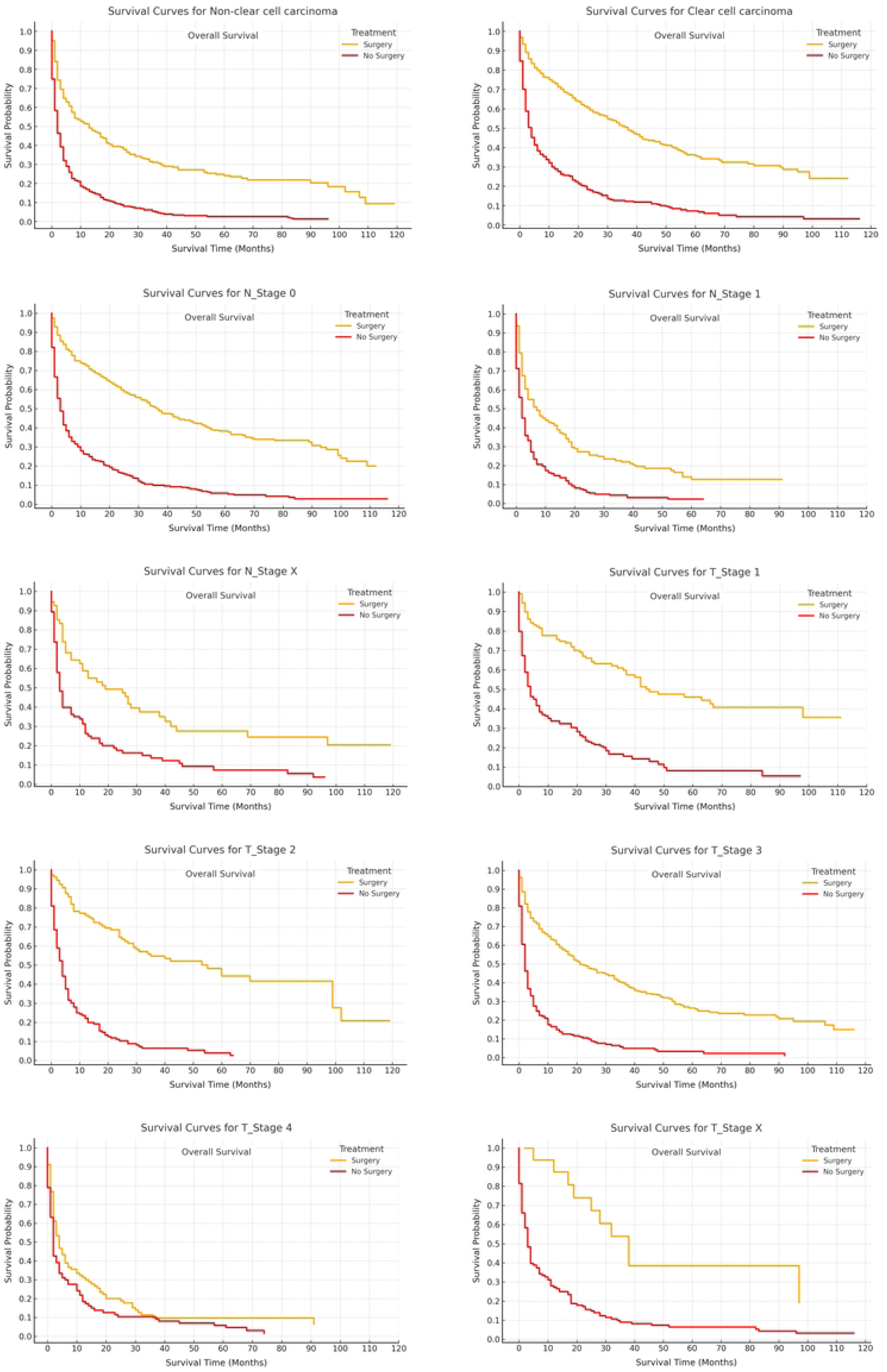
Overall survival stratified by treatment modalities.

### Analysis of the cumulative incidence of competitive risk

As illustrated in Figure 5, the cumulative incidence curve for the surgery group ascends more gradually compared to the non-surgery group, suggesting that surgical intervention may mitigate the cumulative incidence of adverse events. The divergence between the two groups becomes increasingly pronounced over time, underscoring the potential protective effect of surgical treatment during long-term follow-up.

**Figure 5.**
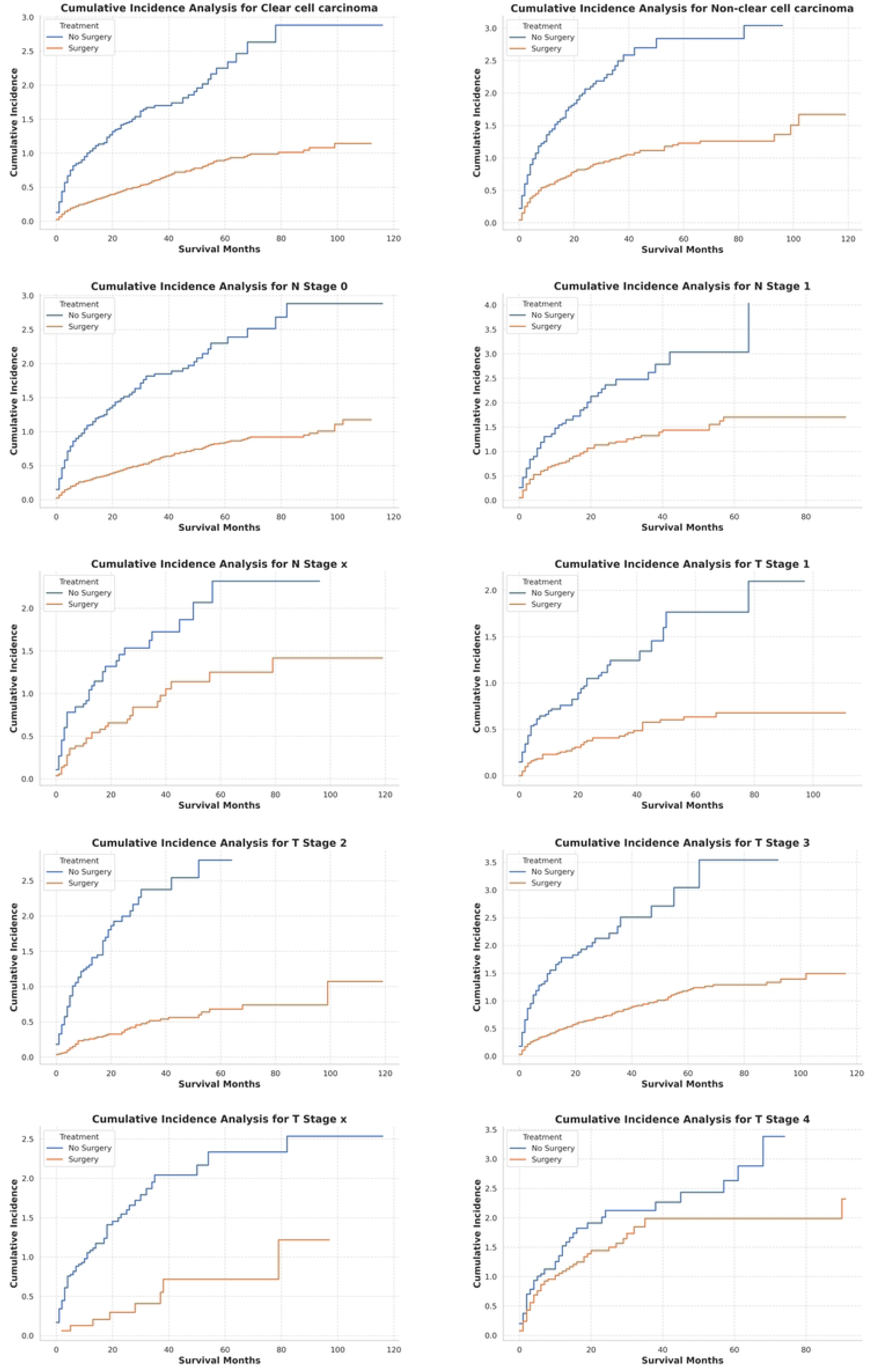
Analysis of the cumulative incidence of competing risks among different treatment modalities.

Moreover, as illustrated in Figure 6, subgroup analyses examining the cumulative incidence of competing risks across various categories (T1–Tx, N0–Nx, clear cell, and non-clear cell histologies) consistently revealed a more gradual increase in the surgery cohort compared to the non-surgery cohort. This pattern suggests a diminished cumulative incidence of adverse events associated with surgical intervention. The widening gap between the treatment groups over time further underscores the potential long-term protective effects of surgery, spanning across distinct tumor stages, nodal statuses, and histological subtypes.

**Figure 6.**
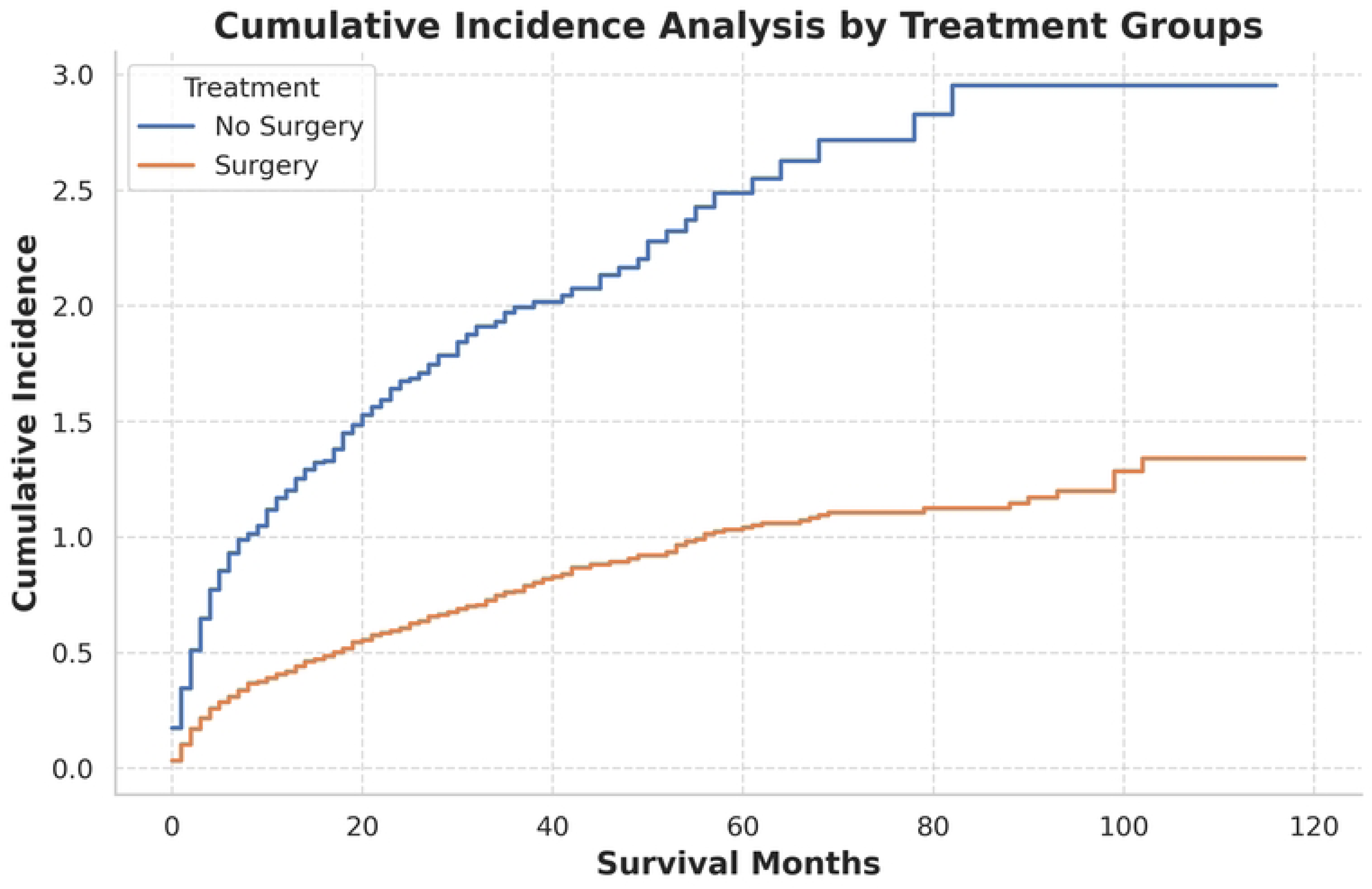
Analysis of the cumulative incidence of competing risks among different subgroup.

## Discussion

This study thoroughly assessed the survival advantages of CN in patients with mRCC by utilizing data from the SEER database. By offering real-world, population-based evidence, our results emphasize the critical role of CN in refining treatment approaches for mRCC. The results revealed significant reductions in both overall mortality and RCC-specific mortality, with consistent survival advantages observed across diverse subgroups. These findings validate the integration of CN within modern multimodal therapeutic frameworks, particularly in combination with targeted and immune therapies, while also providing a foundation for future prospective studies to refine its clinical application.

We demonstrated that CN is associated with significantly improved overall survival (OS) and RCC-specific survival RCC-SS in patients with mRCC. By analyzing a large, population-based cohort from the SEER database and employing propensity score matching to minimize selection bias, we found that CN resulted in a 71% reduction in all-cause mortality risk and a 71% reduction in RCC-specific mortality risk compared to non-surgical management. These survival benefits were consistently observed across various subgroups categorized by age, sex, race, tumor grade, histologic subtype, T stage, and N stage. Additionally, Kaplan-Meier survival curves and analyses of cumulative incidence in the context of competing risks further validated that CN not only extends survival but also diminishes the cumulative incidence of RCC-related mortality over time.

Our multivariate analysis further highlighted the magnitude of CN’s impact. CN was associated with a 71% reduction in all-cause mortality (HR = 0.29, 95% CI = 0.25–0.33, p < 0.0001) and a 71% reduction in RCC-specific mortality (HR = 0.29, 95% CI = 0.25–0.34, p < 0.0001). Survival benefits were consistent across multiple subgroups, demonstrating improved 1-, 3-, and 5-year survival rates in the CN group compared to the non-surgical group. For instance, the 5-year OS rate was 31.50% in the CN group versus only 3.56% in the non-surgical group (p < 0.0001). These robust statistical findings substantiate the critical role of CN in appropriately selected patients.

Stratified analysis lends further biological plausibility to these findings. Patients with lower-grade tumors (e.g., G2: HR = 0.17, p < 0.0001) and early-stage tumors (e.g., T1: HR = 0.34, p < 0.0001) derived the greatest survival benefits, likely due to reduced metastatic burden and better surgical tolerance. However, even patients with advanced-stage tumors (e.g., T4: HR = 0.70, p = 0.0291) or high-grade disease showed significant survival improvements, suggesting that CN may attenuate systemic disease progression even in aggressive cases. Additionally, subgroup analyses by demographic factors demonstrated consistent benefits across age, sex, and race, reinforcing the generalizability of these results. Collectively, these findings underscore the rationale for integrating CN into multimodal treatment paradigms, particularly alongside modern systemic therapies.

The survival advantage conferred by CN likely stems from its ability to reduce tumor burden, modulate the host immune microenvironment, and synergize with systemic therapies(3, 11, 14). By excising the primary tumor, CN may mitigate the immunosuppressive effects of pro-inflammatory and immunosuppressive factors, such as VEGF and TGF-β(15, 16). Additionally, CN may enhance patient responsiveness to immune checkpoint inhibitors and targeted therapies(17, 18). This benefit is particularly critical in the current era of systemic treatment advances(19–21). Our study further confirms that CN significantly prolongs survival in specific patient subgroups, such as those with early-stage and low-grade tumors, underscoring its pivotal role within the multimodal treatment framework.

Our findings align with and extend previous key studies highlighting the survival benefits of CN in mRCC(22). A retrospective study demonstrated significantly improved 5-year OS and RCC-SS in mRCC patients undergoing CN (HR = 0.29, *p* < 0.0001), consistent with our multivariate analysis(22). Similarly, a systematic meta-analysis confirmed that CN combined with targeted therapies significantly enhances survival, reaffirming its role in multimodal treatment strategies(19). Moreover, the study by Mejean et al.(11) underscored the importance of CN, particularly when integrated with targeted therapies, as a cornerstone of the modern therapeutic framework Consistent with our findings, Massari et al.’s(23) meta-analysis revealed that patients with low-grade (G2) and low-stage (T1) tumors derive the greatest benefit, likely attributable to better surgical tolerance and reduced tumor burden.

However, the findings of the CARMENA and SURTIME trials diverge from our results, as these randomized controlled studies reported limited survival benefits of CN in high-risk patients, such as those with extensive metastatic burden or poor performance status(11, 12). For example, the SURTIME trial conducted by Bex et al.(12) suggested that CN may not confer significant survival advantages in the context of advanced systemic therapies, indicating that its efficacy might be constrained in these subgroups. These discrepancies emphasize the need for further prospective studies to better delineate the role of CN in diverse clinical contexts.

Despite its strengths, this study is not without limitations. The retrospective design and reliance on SEER data inherently introduce potential biases and unmeasured confounders, even with propensity score matching(24). Furthermore, the absence of data on modern systemic therapies, such as tyrosine kinase inhibitors (TKIs) and immune checkpoint inhibitors (ICIs), precludes an analysis of their interaction with CN(25). Additionally, the lack of granular clinical information, such as patient performance status, comorbidities, and metastatic burden, may limit the applicability of our findings to specific patient subgroups(26). Future research should focus on integrating CN with emerging therapies, identifying predictive biomarkers of treatment response, and leveraging advanced analytics to optimize patient selection. These efforts are essential to refine multimodal treatment strategies and improve outcomes for mRCC patients.

In conclusion, our study provides robust real-world evidence for the survival benefits of CN in mRCC, underscoring its critical role within multimodal treatment frameworks. Addressing current limitations and exploring the synergy between CN and systemic therapies will enable the development of patient-specific strategies, maximizing therapeutic outcomes and advancing the field of mRCC management.

## Conclusion

CN significantly improves survival in metastatic RCC patients, with consistent benefits across subgroups. This study highlights CN’s critical role in multimodal treatment strategies and provides valuable evidence for its integration with systemic therapies, supporting personalized approaches for optimized patient outcomes.

## Data Availability

All data produced in the present study are available upon reasonable request to the authors,

https://seer.cancer.gov/

## Acknowledgments

We extend our gratitude to the staff of the SEER Database for providing the essential resources that significantly contributed to this study. Additionally, we express our appreciation to the editorial team and reviewers for their insightful and constructive comments, which have substantially enhanced the quality of this manuscript.

